# Alien limb in the corticobasal syndrome: phenomenological characteristics and relationship to apraxia

**DOI:** 10.1101/19013607

**Authors:** David J. Lewis-Smith, Noham Wolpe, Boyd C.P. Ghosh, James B. Rowe

**Author notes:** Corresponding author: Dr Noham Wolpe, Department of Clinical Neurosciences, University of Cambridge, Herchel Smith Building, Cambridge CB2 0SZ, UK. These authors contributed equally to this work (David J. Lewis-Smith, Noham Wolpe).

## Abstract

Alien limb refers to movements that seem purposeful but are independent of patients’ reported intentions. Alien limb often co-occurs with apraxia in the corticobasal syndrome, and anatomical and phenomenological comparisons have led to the suggestion that alien limb and apraxia may be causally related as failures of goal-directed movements. Here, we characterised the nature of alien limb symptoms in patients with the corticobasal syndrome (n=30) and their relationship to limb apraxia. Twenty-five patients with progressive supranuclear palsy Richardson syndrome served as a disease-control group. Structured examinations of praxis, motor function, cognition and alien limb were undertaken in patients attending a regional specialist clinic. Twenty-eight patients with corticobasal syndrome (93%) demonstrated significant apraxia and this was often asymmetrical, with the left hand preferentially affected in 77% of patients. Moreover, 83% reported one or more symptoms consistent with alien limb. The range of these phenomena was broad, including changes in the sense of ownership and control as well as unwanted movements. Regression analyses showed no significant association between the severity of limb apraxia and both the occurrence of an alien limb and the number of alien limb phenomena reported. Bayesian estimation showed a low probability for a positive association between alien limb and apraxia, suggesting that alien limb phenomena are not likely to be related to severe apraxia. Our results shed light on the phenomenology of these disabling and as yet untreatable clinical features, with relevance to theoretical models of voluntary action.

## INTRODUCTION

Alien limb phenomena are a heterogeneous group of behaviours in which one or more of a patient’s limbs, usually an arm, behaves in a manner that appears purposeful or semi-purposeful but is independent of the patient’s reported intentions [1–5]. Patients’ reactions to the unwanted motor behaviours are variable, including lack of awareness, surprise, frustration or denial of ownership of the limb itself [6]. Alien limb motor phenomena have been divided into two main groups [3]. The first group includes complex, unwilled motor acts, including intermanual conflict, mirror movements, interference, and the pushing aside of the directed limb by the autonomous limb. These movements have been described as often bimanual, and liable to occur within two scenarios: (a) the offending hand is involuntarily recruited to tasks which the patient intends to perform unimanually with the other hand and (b) the offending limb undertakes the incorrect act when desired to act in concert with the other hand. The second group of phenomena include simple, unwilled, quasi-reflex actions. These include autonomous reaching, grasping and utilisation behaviour, automatic limb withdrawal or levitation.

Despite ample clinical and basic research, the aetiology of alien limb remains poorly understood [5]. Case studies of patients with focal brain lesions have implicated frontal brain regions [7]. An anatomically derived classification of alien limb phenomena identified two separate syndromes [8]. One syndrome associated with frontal callosal damage, with or without bilateral frontal involvement, typically presents with intermanual conflict (in which involuntary non-dominant hand activity is precipitated by the internally-evoked actions of the dominant hand). The other is a syndrome seen following damage to the left frontomedial and callosal regions in which the patient’s dominant hand exhibits unintentional reaching and grasping evoked by external stimuli [8]. Within these frontal regions, damage to the pre-supplementary motor area may play a central role in the development of alien limb [9, 10].

Alien limb is a common clinical feature of the corticobasal syndrome. Corticobasal syndrome classically arises from the specific pathology termed ‘corticobasal degeneration’ [11], which is associated with aggregation of hyperphosphorylated 4-repeat tau protein [12]. Corticobasal syndrome can also arise from other neuropathologies, such as Alzheimer type pathology [13–15], Creutzfeldt-Jakob disease [14] or the cumulative effects of cerebrovascular disease [16]. The corticobasal syndrome can present with subcortical motor features (including akinetic-rigidity, dystonia and myoclonus) or cortical features (including visuospatial and deficits, non-fluency and cortical sensory loss) [17, 18]. The current international consensus clinical diagnostic criteria for corticobasal syndrome include alien limb [19], as have earlier diagnostic criteria [20, 21].

Another disorder of the sensorimotor system that is included in the diagnostic criteria of, and common in the corticobasal syndrome is called ‘apraxia’ [7, 9, 10, 22]. Apraxia may be defined as a disorder of “the execution of learned movement which cannot be accounted for either by weakness, incoordination or sensory loss, or by incomprehension of or inattention to commands” [23]. It is assessed through the pantomiming of actions (performing an action on request without example) and imitation of gestures [18]. It can result from focal lesions, classically in the corpus callosum and dominant hemisphere [23, 24], as well as neurodegenerative disorders such as corticobasal syndrome [18, 19]. Limb apraxia is usually considered in three forms, often called ‘ideational’, ‘ideomotor’, and ‘limb-kinetic apraxia’ [18, 25]. Ideational apraxia results from a conceptual deficit of the desired action. Ideomotor apraxia is a failure to convert the concept of the action into a motor programme to execute the necessary movements; it usually manifests itself through spatiotemporal errors. Limb-kinetic apraxia describes reduced dexterity predominantly affecting fine movements through incoordination of the fingers, beyond that attributable to elementary motor disorders such as pyramidal dysfunction, rigidity, ataxia, tremor or dystonia [26]. Ideomotor and limb-kinetic apraxias appear to be the most common and may co-exist in the corticobasal syndrome [26], although isolated examination of apraxia in patients with co-existent rigidity, dystonia or tremor is often difficult [18, 19].

In addition to their co-occurrence in the corticobasal syndrome, limb apraxia and alien limb syndrome are both disorders of complex sensorimotor function with some similarities. First, shared anatomic substrates have been implicated through association with lesions either directly affecting the supplementary motor, pre-frontal and parietal cortices or causing their disconnection [7, 9, 10, 27, 28]. Second, classical alien behaviours were phenomenologically described as ‘diagonistic dyspraxia’ [27] and ‘magnetic apraxia’ [29]. Third, in patients with corticobasal syndrome, both apraxia severity and alien limb symptoms have been associated with an implicit measure of the sense of agency [10]. However, there are clear differences between these clinical entities. Apraxia describes aberrant voluntary movement whereas the anarchic action of an alien limb is experienced by the patient as involuntary. An alien limb is associated with a sense of foreignness and loss of agency, which is not characteristic of apraxia.

Here, we investigated the phenomenological nature of alien limb symptoms in the corticobasal syndrome and their possible link with apraxia. We performed structured examinations of praxis, motor function, cognition and alien limb in patients with corticobasal syndrome. We summarised the rate of different alien limb symptoms. We then used Bayesian analyses to compare two alternate hypotheses: (i) that alien limb phenomena and apraxia are related to each other versus (ii) that alien limb phenomena are not related, in that there is no association between alien limb phenomena and the severity of apraxia. This evidence-based null result would suggest differences in terms of their functional anatomy and physiology despite co-occurrence in the corticobasal syndrome.

## METHODS

Participants were recruited from a specialist regional neurology clinic for cognitive and movement disorders during a 40-month period, serving a population of approximately one and a half million. Patients met the diagnosis of probable corticobasal syndrome by clinical criteria confirmed at the time of assessment rather than first presentation [17], and were later re-diagnosed under the revised consensus criteria [19], as probable corticobasal syndrome with either probable or definite corticobasal degeneration. Magnetic resonance imaging in each case had not indicated alternative diagnoses. The age, sex and premorbid handedness of participants are summarised in Table 1. Twenty-five patients with possible or probable Progressive Supranuclear Palsy Richardson’s syndrome (PSP-RS) were recruited as a disease control group under the former MINDS-SPSP criteria for PSP [30]. Unlike the current MDS criteria for PSP [31], the former criteria excluded overlap syndromes such as PSP-corticobasal syndrome. The research was carried out in accordance with guidelines and regulations approved by the Cambridgeshire 2 Research Ethics Committee (now ‘East of England – Cambridge Central’), who approved the experimental protocols. All participants gave full, informed, written consent before the experiment.

**Table 1:** Demographic, neuropsychological and behavioural data. (PSP-RS=Progressive Supranuclear Palsy Richardson Syndrome; CBS=Corticobasal Syndrome; ACE-R= Addenbrooke’s Cognitive Examination Revised; MMSE = Mini-mental State Examination; FAB = Frontal Assessment Battery; UPDRS-III = Unified Parkinson’s Disease Rating Scale (Part III motor subscale); p-values < 0.05 were considered statistically significant)

|  |  | PSP-RS |  |  | CBS |  |  | p-value |
| --- | --- | --- | --- | --- | --- | --- | --- | --- |
| | Units | Mean/% | SD | N | Mean/% | SD | N | (t-test, $\chi^2$) |
| *Demography* |  |  |  |  |  |  |  |  |
| Age | years | 70.6 | 7.9 | 25 | 70.3 | 9.1 | 30 | ns |
| Sex | %male | 60 |  | 25 | 37 |  | 30 | ns |
| Handedness | % R | 96 |  | 25 | 97 |  | 30 | ns |
| *Neuropsychology* |  |  |  |  |  |  |  |  |
| ACE-R Total | /100 | 79.1 | 10.4 | 25 | 70.0 | 13.7 | 29 | 0.008 |
| -Attention & Orientation | /18 | 16.4 | 1.8 | 25 | 15.0 | 2.7 | 29 | 0.028 |
| -Memory | /26 | 21.1 | 4.3 | 25 | 17.2 | 4.7 | 29 | 0.002 |
| -Verbal Fluency | /14 | 5.6 | 3.2 | 25 | 7.6 | 2.9 | 29 | 0.027 |
| -Language | /26 | 23.0 | 2.3 | 25 | 21.9 | 2.5 | 29 | ns |
| -Visuospatial | /16 | 13.4 | 4.3 | 25 | 8.4 | 4.5 | 29 | <0.001 |
| MMSE | /30 | 26.4 | 2.8 | 25 | 23.1 | 4.5 | 30 | 0.002 |
| FAB | /18 | 10.9 | 4.3 | 25 | 10.4 | 4.7 | 28 | ns |
| *Motor examination* |  |  |  |  |  |  |  |  |
| UPDRS-III | /108 | 32.2 | 13.3 | 25 | 24.7 | 9.9 | 19 | 0.038 |
| Praxis Total limb | /20 | 19.1 | 2.1 | 24 | 9.3 | 5.2 | 30 | <0.001 |
| -Transitive mime | /6 | 5.7 | 1.0 | 24 | 3.0 | 2.0 | 30 | <0.001 |
| -Intransitive mime | /6 | 5.9 | 0.4 | 24 | 3.7 | 1.9 | 30 | <0.001 |
| -Unilateral imitation | /6 | 5.7 | 0.9 | 24 | 2.2 | 1.8 | 30 | <0.001 |
| -Bilateral imitation | /2 | 1.8 | 0.6 | 24 | 0.4 | 0.7 | 30 | <0.001 |
| -Orofacial | /2 | 2.00 | 0.0 | 22 | 1.79 | 0.6 | 28 | ns |
| *Alien limb questionnaire* | /13 | 0.08 | 0.4 | 25 | 4.23 | 3.1 | 30 | <0.001 |

The cognitive assessment included neuropsychological evaluation with the Revised Addenbrooke’s Cognitive Examination [32, 33], the Mini Mental State Examination [34], and the Frontal Assessment Battery [35]. Motor function was evaluated using part III of the Unified Parkinson’s Disease Rating Scale [36] and recording of myoclonus, mirror movements, dystonias and dyskinesias. On our assessment of apraxia, we focused on a tailored examination scored according to the broad definition of apraxia without attempting to separate subtypes because of the inherent difficulties of clinical differentiation in a complex movement disorder. Praxis was assessed using a structured bilateral 5-part examination. This included the miming of 3 transitive and 3 intransitive unilateral representational actions, testing each in both the left and right hands separately (6 left, 6 right, instructed verbally), and the imitation of 3 unilateral non-representational hand configurations (3 left, 3 right, gestures demonstrated by the examiner) and the imitation of 2 bimanual hand configurations. Orofacial mime (to mime a kiss and yawn, instructed verbally) was also assessed. Each of the 20 actions was scored 1 in the absence of apraxic error and 0 if apraxic errors were observed. For the association with alien limb (below), we focused on limb praxis score (range: 1-18).

We assessed the presence of alien limb phenomena using a screening questionnaire to guide a structured history from both the patient and an accompanying relative or carer who could corroborate and prompt recall of specific events. This approach was chosen, as alien limb events are intermittent and hard to provoke (unlike apraxia), making it necessary to rely on recalled events. All of our corticobasal syndrome patients had the mental capacity to report their symptoms, and we sought to focus on the patient’s own experience of agency which is crucial for characterising and diagnosing alien limb [2]. The questionnaire included a set of 13 possible experiences occurring within the past six months. The questions addressed different forms of alien limb phenomena: autonomous and disobedient limb movement, the interference of tasks, the sense of control and ownership and other sensations relating to the limb. The questions were: 1) Does your hand copy the other hand on its own?* 2) Does your hand ever float up in the air on its own?* 3) Do you prefer to hold your hand with the other hand? 4) Does your hand *feel* fidgety or restless to you? (emphasis on the feel) 5) Does your hand sometimes reach or touch things without you intending it to?* 6) Does your hand touch your face? 7) Do you prefer to keep your hand in your pocket? 8) Does your hand ever try to stop the ’good’ hand doing what you want it to do?* 9) Does your hand ever reach or touch things even when you want it *not* to?* 10) Do you ever wish your hand would go away? 11) Does your hand sometimes feel that does not belong to you?* 12) Does your hand feel like it belongs to somebody else? 13) Does it feel as though someone else is controlling your hand? The six questions marked with an asterisk refer to phenomena which we considered most specific to alien limb, including anarchic hand phenomena, intermanual conflict and loss of the sense of ownership. We also completed the structured praxis examination and alien limb questionnaire with the PSP-RS cohort for validation.

Logistic and linear regression analyses were used to test for an association between limb apraxia severity (measured through the structured praxis examination) and alien limb. Our main analyses focused on the specific alien limb symptoms reported by patients. Age and ACE-R scores were included as covariates of no interest. All variables were z-score scaled before entered to the regression analyses. Statistical analyses were performed in R [37]. Bayesian estimation of regression coefficients and calculation of Bayes factor were performed with the ‘brms’ package, with priors set to default weakly informative priors [38]. All plots were generated with ggplot2 package [39].

## RESULTS

Patient demographics and clinical information are summarised in Table 1. Limb praxis scores were incomplete for one patient with corticobasal syndrome and another with PSP-RS. All patients completed the alien limb questionnaire.

### Praxis Scores

Apraxia was very common in our corticobasal syndrome cohort, with only 1/30 patients demonstrating normal praxis. In corticobasal syndrome, praxis scores ranged between 0/20 (inability to perform any of the required movements) and 20/20 (no deficit) (Fig. 1A). As shown in Table 1, patients with corticobasal syndrome were significantly apraxic in all limb movements tested, resulting in mean total limb praxis score of 9.3/20 (SD 5.2). Of the 29 patients with apraxia, the left hand was preferentially affected in 23 patients; the right hand in four and symmetrical in two patients. The only left-handed corticobasal syndrome patient exhibited predominantly left-sided apraxia. By contrast, most patients with PSP-RS did not have apraxia. Seventeen of twenty-four patients with PSP-RS scored 20/20 (no deficit) and three more scored 19/20.

**Figure 1.**
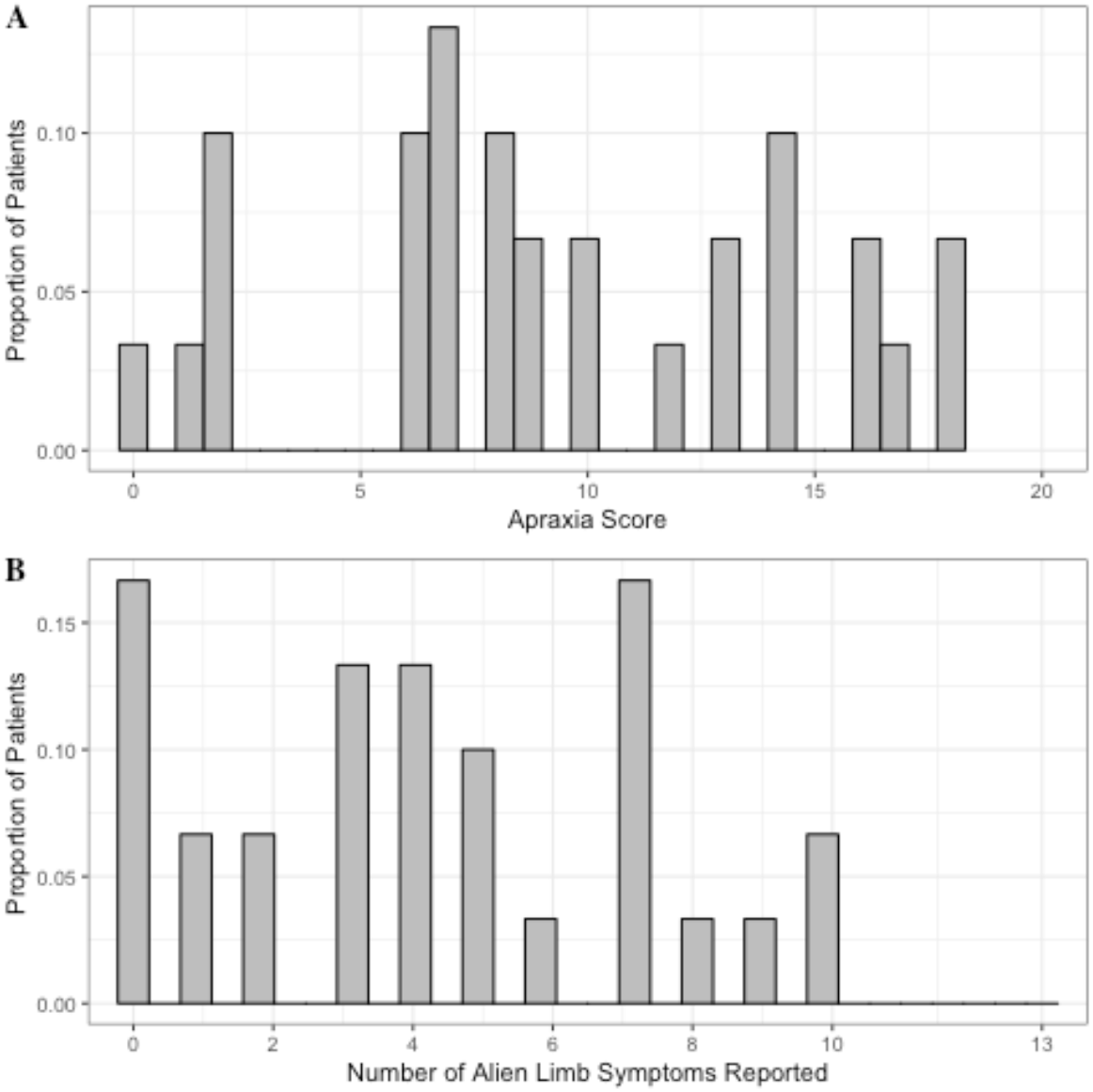
A) The distribution of apraxia score in corticobasal syndrome patients. Lower scores indicate fewer movements successfully performed (that is – more abnormal behaviour). B) Same as (A) but for the overall number of alien limb phenomena reported by patients (higher number indicates more alien limb symptoms reported).

### Alien limb phenomena

Alien limb phenomena were reported in the majority of corticobasal syndrome patients (Fig. 1B) but only one person with PSP-RS (who reported only the non-specific symptom that he had a fidgety hand that he tended to hold). Further analyses are restricted to the corticobasal syndrome group.

Overall, 83% of the patients with corticobasal syndrome reported at least one symptom of alien limb, and 72% reported at least one of the more specific symptoms. These specific symptoms included motor phenomena and whether the patient feels that their limb belongs to them (see Methods). Each individual item received affirmative responses from some patients and their frequency is summarised in Figure 2. Common responses were of the tendency to hold the offending hand with the better hand (53%), for unwilled arm levitation (50%), and the sensation that the limb was not theirs (50%). Patients rarely reported preferring to keep their *bad* hand restricted in a pocket (20%), the *bad* hand blocking the *good* hand (intermanual interference) (17%), the sensation that the limb belonged to somebody else (17%) or that the limb was under the control of another agent (17%). Four of the six patients reporting either of these last two symptoms reported both. We confirmed there was no association between cognitive impairments, measured with the ACE-R, and both the number of specific (Spearman’s rho = 0.022, *p* = 0.909) and overall (Spearman’s rho = 0.202, *p* = 0.294) alien limb phenomena reported.

**Figure 2.**
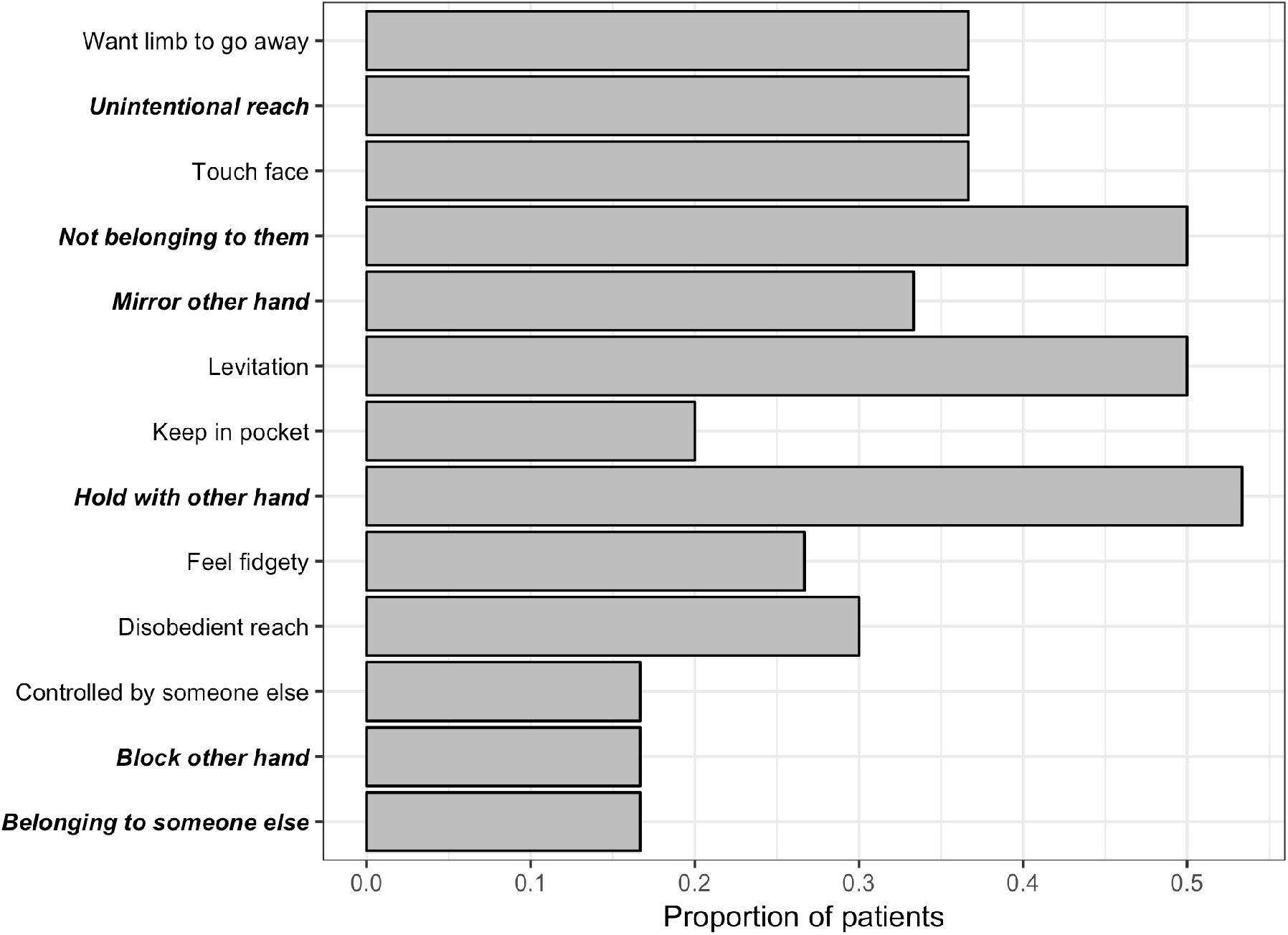
The prevalence of each alien limb symptom in corticobasal syndrome patients. More specific alien limb symptoms in ***bold-italics***. Symptoms ordered alphabetically.

### Association between alien limb and apraxia

We investigated a possible association between alien limb phenomena and limb apraxia in corticobasal syndrome (Fig. 3A), while accounting for age and ACE-R score. First, we conducted a logistic regression analysis to test whether apraxia severity was associated with the occurrence of alien limb in patients. There was no association between praxis score and the emergence of at least one of the specific alien limb symptoms in patients (*z* = -0.726, *p* = 0.468; *OR* = 0.922, 95% *CI* = 0.727-1.142). The results did not change when we looked at the occurrence of at least one of any of the items in the alien limb questionnaire items (*z* = -0.052, *p* = 0.958; *OR* = 0.992, 95% *CI* = 0.729-1.327).

**Figure 3.**
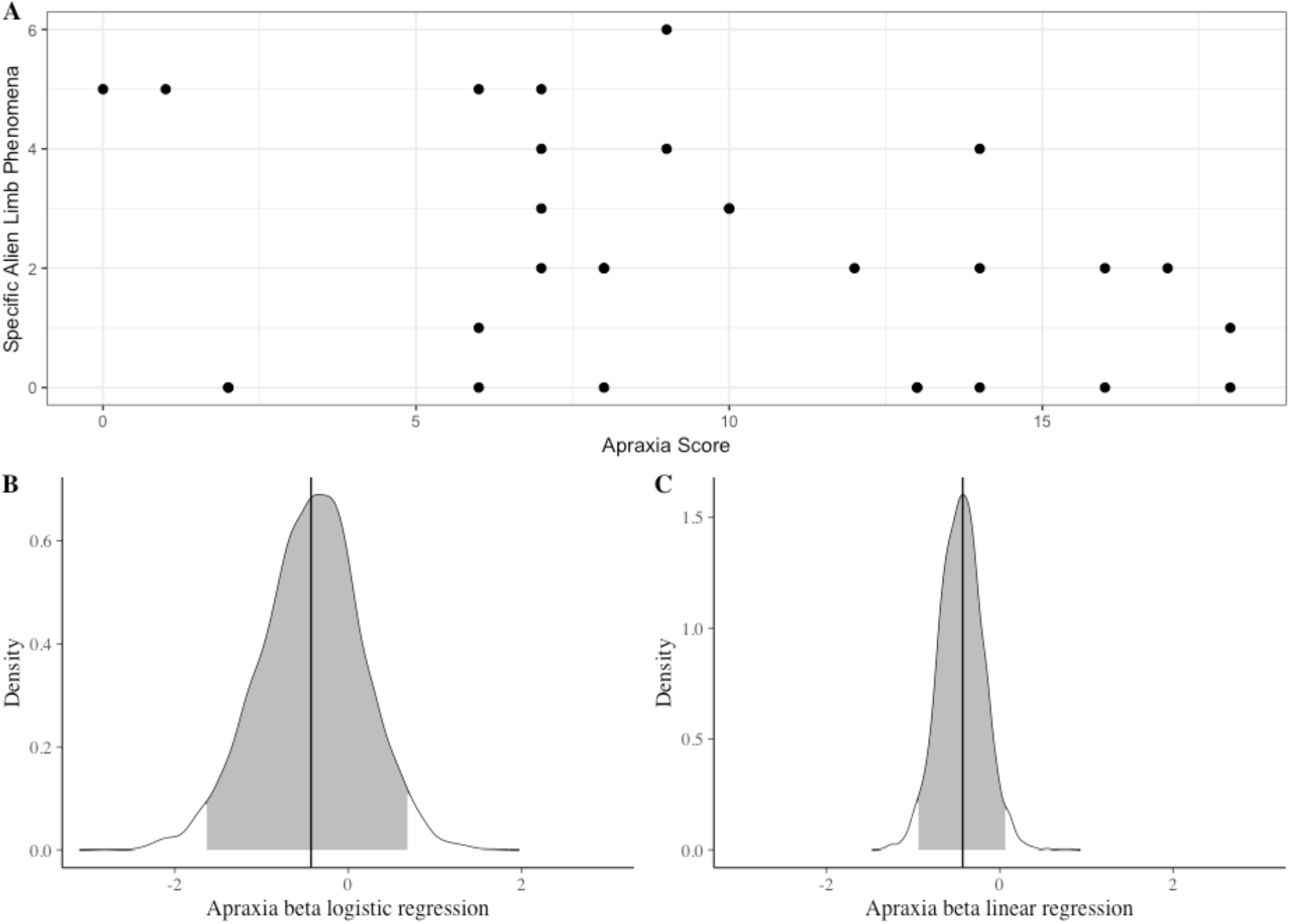
Alien limb and apraxia. A) The number of alien limb symptoms reported by patients plotted against apraxia severity scores. B) Probability density function for the beta coefficient for apraxia predicting the occurrence of at least one of the specific alien limb symptoms in the logistic regression analysis, estimated with a Bayesian model fit. Vertical black line indicates the mode. Grey fill indicates 95% credible interval. C) Same as (B) but for the beta coefficient of apraxia predicting the total number of specific alien limb symptoms in the linear regression analysis.

We performed a Bayesian estimation of the logistic regression coefficients for predicting the occurrence of at least one of the specific alien limb symptom (Fig. 3B). This analysis revealed that the probability for increased risk of developing at least one of the specific alien limb phenomena with increasing apraxia severity was 0.225. Model comparison showed that a logistic regression model that did not include apraxia severity as an independent variable was favoured over a model that did, with a Bayesian factor of 3.68 (moderate evidence in favour of the null).

Next, we examined the phenomenological association between alien limb and apraxia by performing a linear regression analyses on praxis score predicting the number of alien limb symptoms (Fig. 3B). No such association emerged, both when looking at the number of specific alien limb phenomena reported (*beta* = - 0.415, *p* = 0.078) and the overall number of phenomena reported (*beta* = -0.265, *p* = 0.240). A Bayesian estimation of the regression coefficients (Fig. 3C) showed that the probability of apraxia severity positively predicting the number of specific alien limb phenomena reported (i.e. *beta* > 0) is 0.037. Model comparison showed that a linear regression model that did not include apraxia severity as an independent variable was strongly favoured over a model that did with a Bayesian factor of 689.73 (very strong evidence, by conventional thresholds).

For completeness, we considered a non-linear relationship between alien limb and apraxia by performing non-parametric, ranked partial correlation analyses. No correlation was found between apraxia severity and the total (Spearman’s rho = -0.253, *p* = 0.194) or specific (Spearman’s rho = -0.358, *p* = 0.061) alien limb phenomena reported.

As most of the patients suffered from asymmetric apraxia, we completed the analyses with a Bayesian estimation of regression coefficients with lateralised apraxia score instead of total limb apraxia score. This revealed that the probability for a positive association between unilateral apraxia severity and occurrence of alien limb was 0.019 and 0.039 (specific and any alien limb symptom, respectively). Similarly, the probability for a positive association between unilateral apraxia severity and number of alien limb symptoms reported was 0.006 and 0.015 (specific and all alien limb symptoms, respectively).

## DISCUSSION

The principal results from this study are although both alien limb and apraxia are common in corticobasal syndrome, severe apraxia does not predict the occurrence of alien limb and there was no association between the number of alien limb phenomena reported and apraxia severity. These results have the caveat that our data emphasise the qualitative features of alien limb phenomena reported by patients, rather than their frequency or severity. We used a structured assessment of both apraxia and alien limb syndrome in order to formalise the assessment of these disorders for analysis and found them to be robust to the presence of another severe extra-pyramidal disorders with involuntary movements: PSP-RS. Whilst our semi-quantative analysis did not address each form of apraxia separately, the composite limb apraxia score included imitation and mime tasks. These are sensitive to both ideomotor and limb-kinetic apraxia and also in the case of mime, ideational apraxia [18]. We first consider the implications of our results for the understanding of alien limb phenomena and apraxia, before considering the relationship between these clinical entities.

### Alien limb phenomena

Alien limb phenomena were common in our patients with corticobasal syndrome, with 70% reporting at least one of the six specific phenomena (copying, flotation, unintentional reach, oppositional reach, interference, sense of belonging). In our patients, the actions made by alien limbs appeared semi-purposeful and not perseverative as those described due to other aetiologies [3, 6]. Moreover, none of our patients demonstrated self-destructive behaviours as those depicted in case reports arising from other aetiologies [40, 41], suggesting that such phenomena are uncommon in corticobasal syndrome.

A prominent view of alien limb stresses the need for both ‘foreignness’ and involuntary motor activity [2]. On this view, these features are necessary in order to differentiate alien limb phenomena from purely motor disorders, such as dystonia, grasp reflex, athetosis, hemiballismus, hemiataxia and utilisation behaviour, as well as from sensory neglect syndromes [2]. Half of our patients met this definition of an alien hand, experiencing a loss of sense of ownership – that is, the feeling that their hand did not belong to them, together with unwanted movements (i.e. motor phenomena). By contrast, six other patients could be considered to have an ‘anarchic hand syndrome’, as they reported specific motor phenomena while maintaining a sense of ownership [42]. Each of the 15 patients who reported the sensation that their limb did not belong to them also reported at least one specific motor phenomenon. This finding suggests that unwilled motor behaviour may be necessary, but not sufficient for the loss of ownership.

Longitudinal data will be required to investigate whether alien limb results from a two-step process, where a patient first develops an anarchic limb and subsequently loses their sense of ownership. Importantly, while half of our patients reported a feeling of loss of limb ownership, only a third of these reported a sensation of possession or passivity. Therefore, the type of perceived loss of control observed in our patients is distinct from the classic somatic delusions of control which may be experienced by patients with psychosis, and some reported patients with alien limb syndromes of other etiologies [2, 6, 8, 40, 41].

More than a third of our patients reported unintentional reaching movements. Reaching and grasping movements of an alien limb have been suggested to result from ‘exaggerated affordance’, in which motor schema are abnormally disinhibited [43]. These motor plans are considered automatic as they are stimulus (e.g. object-) driven, and in the case of alien limb, a failure to inhibit such motor plans leads to unwanted movements [43].

### Apraxia

Nearly all patients with corticobasal syndrome were found to be apraxic before the degree of akinesia, rigidity, dystonia or myoclonus precluded interpretation of motor deficits as apraxic. 26/29 of the affected patients had significant asymmetry in their apraxia. This is consistent with the common presenting complaint (in corticobasal syndrome) of unilateral ‘clumsiness’ of hand movements. The impact on mime, imitation and tool use was not simply a function of the complexity of the desired action, suggesting the observed motor deficits in corticobasal syndrome are unlikely to be attributable to limb-kinetic apraxia [18, 25]. Our series also found left-sided predominance of asymmetrical apraxia. This is reminiscent of the apraxia from callosal lesions, which has been attributed to interhemispheric disconnection affecting the transfer of motor programmes from the dominant to non-dominant hemisphere [23].

Our test battery for apraxia was based on a simple task description (imitation, intransitive and transitive mime), rather than the inferences of ideational, ideomotor and limb-kinetic subtypes. There are several reasons for this. First, we did not assume that the presentations of apraxia would be the same in a chronic degenerative and distributed disease as in acute, focal surgical or stroke lesions. The complexity of motor deficits arising from combined cortical and basal-ganglia degeneration might obscure the classical dissociations and interpretation of apraxic movements [18, 25]. It has been suggested that ideomotor apraxia is the most common apraxia in corticobasal syndrome [19] while ideational apraxia (revealed by identification and recognition) is less common in corticobasal syndrome [18]. It remains uncertain to what extent the poverty of fine finger movements in corticobasal syndrome is attributable to limb-kinetic apraxia as a true form of apraxia, as opposed to other concurrent pyramidal and extrapyramidal impairments [18, 25]. Moreover, limb-kinetic apraxia in corticobasal syndrome often co-exists with ideomotor apraxia [26]. Hence, we opted for a functional description in terms of the tasks and an aggregate score of the deficits rather than subtyping the apraxias in corticobasal syndrome.

### The relationship between alien limb phenomena and apraxia

Alien limb and apraxia have been both associated with changes in implicit measures of awareness and control of action [10]. Their co-occurrence and shared anatomical substrates suggested that they might be mechanistically related. However, in our cross-sectional cohort of patients with corticobasal syndrome, we found no association between the occurrence or the number of reported alien limb phenomena and limb apraxia severity. Bayesian analyses and model comparison showed that apraxia severity is very unlikely to be a positive predictor of alien limb symptoms. These results do not support the hypothesis that severe apraxia leads to alien limb. Instead, our findings on the phenomenology of alien limb are consistent with a syndrome in which a patient first presents with an involuntary motor disorder and subsequently develops a sensation of ‘foreignness’ or ‘alienness’. The emergence of the motor phenomena of alien limb, even those which have previously been considered *dyspraxic* in nature [27, 29], appears to bear no simple association with the current severity of apraxia.

The clinical and phenomenological dissociation between apraxia and alien limb phenomena have a number of potential explanations, and implications for the theoretical frameworks accounting for both disorders. Firstly, alien limb and apraxia may have overlapping, but not identical underlying brain lesions [7, 22]. Specifically, the pre-supplementary motor area plays a key role within a prefrontal network critical to both alien limb and apraxia, however, the exact localisation of brain lesions within this network may determine the specific manifestation of alien limb, apraxia or both [10]. A disconnection of this prefrontal network from posterior parietal regions that integrate spatio-temporal signals may lead to apraxia [10, 44], whereas damage to internal feedback loops may lead to an inappropriate activation of motor schema that result in alien limb phenomena [10, 43, 45].

Secondly, different pathological processes may selectively affect the functions of a shared network mediating praxis and voluntary movement. Our patients had clinically defined corticobasal syndrome, with neurodegeneration (rather than metabolic or cerebrovascular disease), but pathological heterogeneity is common within this syndrome [13, 15]. It is conceivable that different pathological mechanisms could cause corticobasal syndromes with different motor disorders, although the presence of corticobasal degeneration vs Alzheimer pathology is not directly distinguished by alien limb or apraxia [13, 15].

A third possibility is a complex temporal relationship during disease progression in which apraxia and alien limb phenomena develop out of phase. The relationship between apraxia and alien limb phenomena may then not be evident in a cross-sectional study. For example, as praxis worsens, alien limb phenomena might diminish due to their obscuration by dystonia or akinetic-rigidity [46]. In the three patients with serial praxis examinations and alien limb questionnaires, praxis always deteriorated while alien limb phenomena emerged and sometimes disappeared again over time (data not shown). One might speculate that as the pathways which mediate praxis degenerate in corticobasal syndrome, transient imbalances occur between networks for motor control, attention and awareness which lead to manifestations of alien limb phenomena. The independence of corticobasal alien limb phenomena from apraxia may therefore not simply generalise to cases with other aetiologies, such as brain lesions that concurrently affect regions common to both disorders.

Our study has several limitations. First, unlike the examination of apraxia, our assessment of alien limb symptoms relied on a structured questionnaire. This is because alien limb symptoms are intermittent and hard to provoke in the clinic (unlike apraxia). Responses were corroborated by the carer, and we assessed PSP-RS as a control patient group, which as expected reported no alien limb symptoms. Within the corticobasal syndrome group, certain responses were consistently rare, such as sensation of possession, which could serve as negative controls. Second, we quantified the number of distinct alien limb features, not their frequency or functional impact. In pilot work, patients and their companions found reporting of the frequency of paroxysmal alien phenomena difficult. This may reflect lack of awareness of some episodes, recall bias for episodes that were socially distressing, or the influence of other cognitive and motor features on the functional impact of alien limb. Moreover, we prioritised the description of actual alien limb phenomena, which are not operationalised in the current consensus diagnostic criteria [19]. Third, our cross-sectional study is unable to assess the timeline for the development of apraxia and alien limb, for which longitudinal studies are needed.

## CONCLUSIONS

Alien limb phenomena and apraxia are both common disorders in the corticobasal syndrome. We describe the phenomenology of alien limb in the corticobasal syndrome, as reported by patients and apraxia as observed by clinicians. Our data do not support the interpretation of alien limb phenomena resulting from apraxia in neurodegenerative disease. There might be qualitative differences from the alien limb phenomena and apraxia following focal brain lesions, but such features are less well characterised than in corticobasal syndrome where alien limb and apraxia are both diagnostic features. Longitudinal and quantitative studies are needed to confirm that the alien limb is a transient feature, whereas apraxia declines continuously, independent from one another.

## Data Availability

All data are available upon reasonable request from the corresponding author.

## ACKNOWLEDGEMENTS

DLS was funded by the Guarantors of Brain Entry Fellowship. NW and JBR were supported by the James S. McDonnell Foundation 21st Century Science Initiative, Scholar Award in Understanding Human Cognition. JBR was also supported by the Wellcome Trust [103838] and the Medical Research Council [SUAG/004 RG91365] and the National Institute for Health Research Cambridge Biomedical Research Centre including the Cambridge Brain Bank. BCPG was funded by a MRC Fellowship [G0700503], Guarantors of Brain and the Raymond and Beverley Sackler Trust.

